# CPAP Implementation Strategies for Optimized Neonatal Care: A Scoping Review

**DOI:** 10.1101/2025.09.11.25335579

**Authors:** S. Musungu, K. Dougherty, N. Kundu, C. Miller, C.A. Bohne, L R. Hirschhorn

## Abstract

**Objective:** This scoping review aims to identify change ideas and implementation strategies that support CPAP use for eligible newborns (<2500g, preterm, or with RDS symptoms), and to inform quality improvement efforts in NEST360-supported facilities in sub-Saharan Africa through relevant process measures.

**Introduction:** Continuous Positive Airway Pressure (CPAP) is a well-established and effective treatment for neonates with respiratory distress syndrome (RDS), widely used to prevent respiratory failure in small and sick infants across different settings. Yet, its timely and appropriate use remains inconsistent due to persisting obstacles to access and implementation, particularly in sub-Saharan Africa. Gaps in CPAP coverage related to challenges in device availability, functionality and delivery continue to hinder the provision of optimal care to vulnerable newborns. While the barriers have been widely documented, few literature reviews have systematically explored tangible strategies aimed at overcoming these barriers.

**Inclusion criteria:** This review includes articles involving preterm and term neonates (<28 days) with birth weight <2500g and/or symptoms of respiratory distress syndrome (RDS), thus eligible for CPAP therapy. It focuses on interventions that support timely initiation and appropriate use of CPAP, including implementation strategies and change ideas for quality improvement. Eligible clinical settings include neonatal intensive care units, hospital wards, delivery rooms, and other newborn-focused environments. No geographical restrictions were applied.

Studies will be excluded if they (a) do not center the defined neonatal population, (b) exclusively assess CPAP effectiveness rather than implementation, (c) explore CPAP use for unrelated conditions (e.g., sleep apnea), or (d) described barriers without corresponding solutions.

**Methods:** This scoping review follows the Joanna Briggs Institute methodology and uses the Population–Concept–Context framework to guide inclusion criteria. This protocol is published a priori in alignment with PROSPERO guidelines. Searches were conducted in MEDLINE (via PubMed), Embase (via Elsevier), and CINAHL (via EBSCOhost) in August 2025, with no date limits and English-language restriction. The search strategy was peer-reviewed using the PRESS 2015 guideline. Reference lists of included studies will be screened for additional sources.

Article selection will be performed independently by two reviewers using Rayyan, with discrepancies resolved by consensus or a third reviewer. Full-text screening will apply predefined inclusion criteria, and reasons for exclusion will be documented. Data extraction will be conducted using a piloted form developed by the review team, capturing facility and participant characteristics, implementation strategies/change ideas, context, and CPAP-related use outcomes. No critical appraisal will be performed, consistent with JBI guidance on scoping reviews. Results will be presented narratively and in tabular format, with a PRISMA-ScR flow diagram summarizing the selection process.

## Introduction

Despite declining rates globally, neonatal mortality - defined as death occurring within the first 28 days of life-remains disproportionally high in low-resource settings. In 2023, the United Nations Children’s Fund (UNICEF) estimated a neonatal mortality rate of 26 deaths per 1,000 live births in sub-Saharan Africa, making a child born in that region over ten times as likely to die in the first month compared to a child born in a high-income country.^1^ In 2015, the World Health Organization (WHO) introduced Sustainable Development Goal (SDG) Target 3.2 aiming to end preventable deaths of newborns and children under five by 2030. At the time of its adoption, the global neonatal mortality rate stood at approximately 19 deaths per 1,000 live births. By 2023, this figure had declined to 15 deaths per 1,000 live births.^2^ Still, recent projections indicate that over 60 countries are unlikely to meet the SDG target, most of them in sub-Saharan Africa.^2^ In these regions, neonatal mortality rates remain as high as 26 deaths per 1,000 live births, with most fatalities resulting from preventable complications. Achieving this SDG target will require identifying actionable solutions and evidence-based interventions to improve the quality of small and sick newborn care (SSNC).

Among the leading causes of neonatal mortality, respiratory distress holds particular significance due to its high prevalence and rapid progression.^2^ It contributes to nearly 40 percent of under-five deaths when considered alongside birth complications such as asphyxia, neonatal infections, and congenital anomalies.^1^ Respiratory distress syndrome (RDS) is a severe breathing disorder that often manifests within minutes to hours of birth, typically in premature infants whose lungs may be underdeveloped. However, it is not limited to prematurity; term neonates with low birth weight (under 2500g) due to intrauterine growth restriction or compromised delivery conditions are also at increased risk.^3,4^ Timely intervention is critical, as delayed treatment is strongly associated with adverse outcomes including respiratory failure, multi-organ dysfunction, and death.^5^

WHO recognizes continuous positive airway pressure (CPAP) as a cornerstone for non-invasive treatment of neonatal RDS.^6^ The CPAP device is placed on the nose and functions by delivering a constant distending pressure to the airways, which helps maintain alveolar patency and alleviate RDS symptoms. This intervention stabilizes the lungs and facilitates gas exchange while reducing the infant’s work of breathing.^7^ Although CPAP was first described in 1911, its clinical application for neonatal respiratory distress was reintroduced in 1971 by researchers at the University of California, San Francisco.^7^ Since then, CPAP has been shown to significantly reduce the need for mechanical ventilation, lower the risk for respiratory failure, and overall improve outcomes for at-risk newborns.^8^ However, satisfactory outcomes depends on factors such as timing of initiation, reliable equipment, appropriate patient selection, and sufficient human resources.^9-11^ For example, a study from district hospitals in Malawi found that rigid provider roles contributed to delays in CPAP initiation, underscoring persistent operational barriers in low-resource settings.^10^

Although the clinical effectiveness of CPAP is well-established, its adoption in low-resource settings remains limited, contributing to persistently high rates of preventable neonatal mortality.^11^ The NEST360 alliance was launched to improve the quality and coverage of hospital-based newborn care in Africa. The initiative brings together local and global clinicians, biomedical experts, and public health leaders working with governments in scaling WHO-approved technologies and implementing evidence-based practices across Kenya, Malawi, Nigeria, Tanzania, and more recently, Ethiopia.^12^ As part of this effort, CPAP devices were distributed to NEST360-supported hospitals along with tailored training packages to support their use and maintenance along with training and support for data use to identify challenges and quality improvement. One of the key outcomes monitored by NEST360 is the proportion of eligible newborns who receive CPAP, and subsequent trends in clinical outcomes.

There have been increases in CPAP use across the NEST360-supported sites related to ongoing training and QI, but variation in CPAP coverage rates has been observed across hospitals and countries.^13^ These results reflect challenges to CPAP implementation that have been a focus on prior studies and quality improvement (QI) efforts in similar setting. In addition to ensuring the reliable availability of equipment, common barriers identified include staffing shortage, inefficient training models, lack of bioengineering support for device use and troubleshooting, and other difficulties related to role separation, management and supervision, diagnostic protocols, and communication with caregivers.^9-11^ Building from this work to described barriers to effective CPAP implementation in low-resource settings, this scoping review focuses on implementation strategies and change ideas that have been used to tackle these barriers. An implementation strategy is defined as a bundle of actions taken to enhance adoption, implementation and sustainability of evidence-based interventions.^14^ Change ideas are defined as specific ideas, from research or other organizations, that can be tested to improve a system or process.^15^

### Objectives

This scoping review aims to identify change ideas and implementation strategies employed across clinical settings to support the initiation and use of CPAP for eligible newborns, namely preterm and term neonates (<28 days) with birth weight <2500gor RDS symptoms. The review will also extract commonly reported barriers to CPAP implementation and map them to tested or proposed solutions, assessing the extent to which these interventions have demonstrated success or limitations in overcoming the barriers and improving CPAP delivery. The overarching goal is to synthesize actionable knowledge to inform QI efforts in NEST360-supported countries across sub-Saharan Africa. This includes identifying process measures to evaluate and iteratively refine future implementation activities.

## Review question

What change ideas and implementation strategies have been used to promote the timely initiation and use of CPAP for eligible newborns in clinical settings?

## Inclusion criteria

### Participants

This review will include studies involving small and sick newborns (SSNC) which include premature and term neonates under 28 days of life, with low birthweight (<2500g), or RDS symptoms. Studies will be excluded if they focus on non-neonatal populations, infants older than 28 days, or neonates without documented RDS symptoms or risk factors.

### Concept

The central concept of this review is the use of CPAP therapy for eligible newborns. Included studies will examine implementation strategies and change ideas aimed at optimizing CPAP delivery, accompanied by evidence of success or failure. Studies will be excluded if focused solely on the clinical effectiveness of CPAP for treating RDS, address CPAP use for unrelated conditions (e.g., sleep apnea) or describe implementation barriers without proposing actionable solutions.

### Context

The context of this review includes clinical settings focused on newborn care including neonatal intensive care units (NICUs), standard neonatal units, and other inpatient environments that provide care for infants at risk both in high-income and low- and middle-income countries. Delivery rooms and obstetric care units will be included only if the study emphasizes neonatal care and CPAP use within those settings.

### Types of sources

This scoping review will consider both experimental and quasi-experimental study designs including randomized controlled trials, non-randomized controlled trials, before and after studies and interrupted time-series studies and quality improvement (QI) projects. In addition, analytical observational studies including prospective and retrospective cohort studies, case-control studies and analytical cross-sectional studies will be considered for inclusion. This review will also consider descriptive observational study designs including case series, individual case reports and descriptive cross-sectional studies for inclusion.

In addition, systematic reviews that meet the inclusion criteria will also be considered, depending on the research question.

Purely qualitative studies as well as opinion papers will not be considered for inclusion in this scoping review.

## Methods

This scoping review follows the Joanna Brigg’s scoping review framework.^16^ We used the Population-Concept-Context framework to inform the inclusion criteria and developed an a *priori* protocol based on the PROSPERO guidelines (See Appendix A). The following paper reports the protocol to be followed for the review process, with the aim to limit deviations from the stated strategy.

### Search strategy

The search strategy aims to locate published studies only. A three-step search strategy will be employed in this review. First, an initial limited search of MEDLINE (via PubMed), Embase, and CINAHL (via EBSCOhost) was conducted to identify relevant articles on the topic. The text words contained in the titles and abstracts of retrieved articles, along with the index terms used to describe them, were analyzed to inform the development of a comprehensive search strategy.

A full search strategy was then developed for each selected database, incorporating all identified keywords and index terms. The databases to be searched include MEDLINE (via PubMed), Embase (via Elsevier), and CINAHL (via EBSCOhost). The complete search strategies for each database are provided in Appendix B. The search strategy was peer-reviewed by a librarian using the PRESS 2015 Guideline Explanation and Elaboration.^17^

The reference lists of all included sources of evidence will be screened to identify additional studies. This will include relevant reviews, case studies, cohort studies, and other designs as appropriate. These articles will be screened using the same inclusion criteria outlined in this protocol.

Studies published in languages other than English will be excluded if no translation is available, due to the language capacities of the review team. No date limits will be applied to the search.

### Study/Source of evidence selection

Following the search, all identified citations will be collated and uploaded into EndNote 21 (Clarivate Analytics, PA, USA), and duplicates will be removed. Screening of titles and abstracts will be conducted by two independent reviewers using *Rayyan*, a web-based platform designed to facilitate systematic reviews through collaborative tagging, filtering, and semi-automated screening features.^18^ Potentially relevant sources will be retrieved in full, and their citation details will be imported EndNote 21, and data extraction will be conducted in Excel. The full text of selected citations will be assessed in detail against the inclusion criteria by two independent reviewers. Reasons for exclusion of sources of evidence at full text that do not meet the inclusion criteria will be recorded and reported in the scoping review. Any disagreements that arise between reviewers at any stage of the selection process will be resolved through discussion or consultation with a third independent reviewer. The results of the search and study inclusion process will be reported in full in the final scoping review and presented in a PRISMA-ScR flow diagram.^19^

### Data extraction

Data will be extracted from papers included in the scoping review by two independent reviewers using a data extraction tool developed by the reviewers. This tool was based on the *NEST360/UNICEF Small and Sick Newborn Care (SSNC) Core Components Framework*, which outlines nine interdependent components essential for delivering high-quality, family-centered care: leadership and governance, data systems and quality improvement, referral systems, infrastructure, equipment and commodities, infection prevention and control, linkage to maternal care, post-discharge follow-up, financing, and human resources (Figure 1).^20^

**Figure 1.**
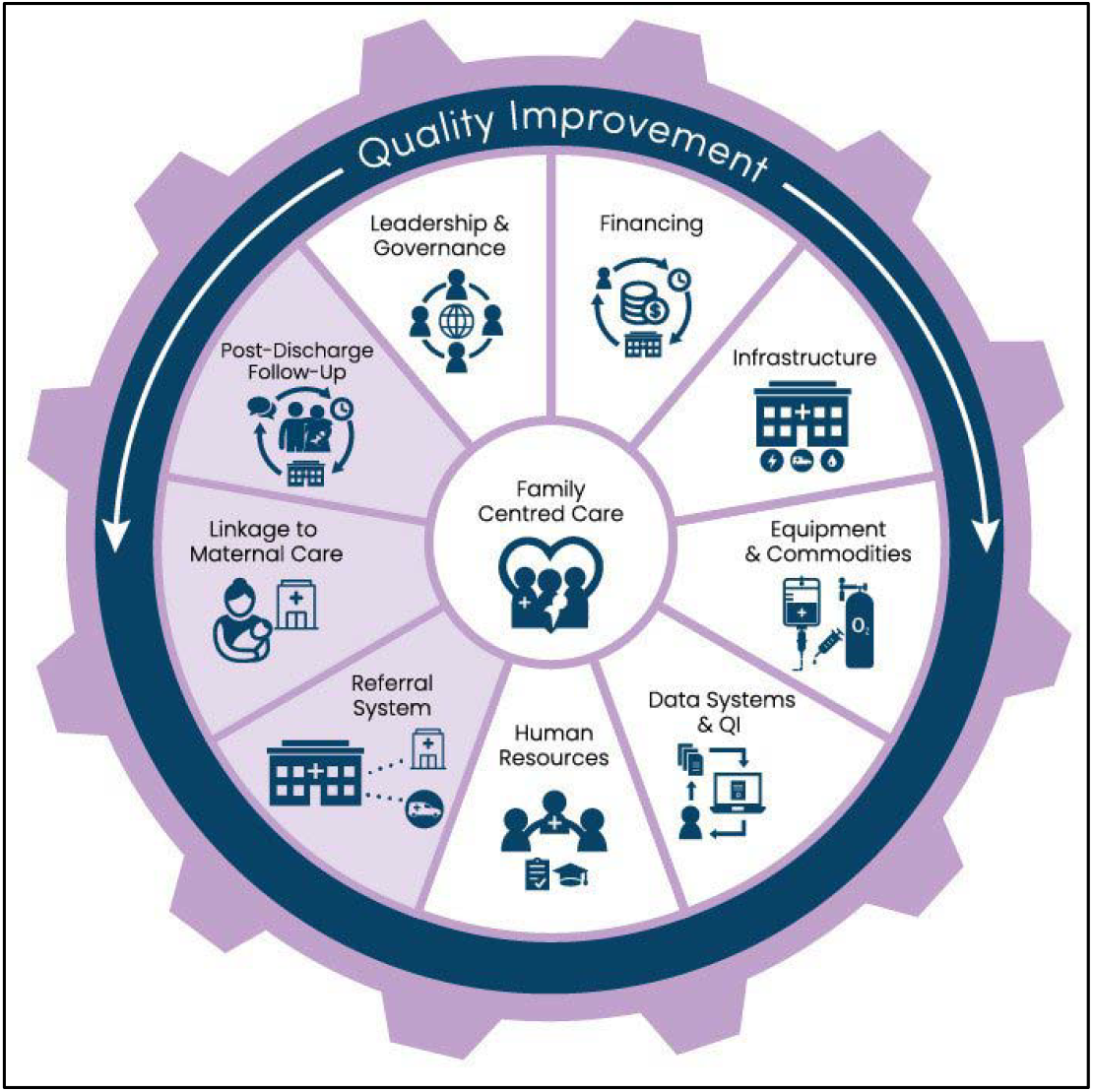
NEST360/UNICEF Core Components for Small and Sick Newborn Care.^20^.

The contingent SSNC Core Components framework assembles experiences, knowledge, and resources that were used to guide the organization of change ideas. The resulting form was adapted to include fields for contextual factors and reported barriers and facilitators. Additionally, Proctors implementation outcomes definitions were used to define the outcomes for identified implementation strategies and change ideas.^21^ A draft version is provided in Appendix C.

Data extraction will be completed in Rayyan. Extracted data will include details about participants, concept, context, study methods, and key findings relevant to the review question. The form will be piloted on a small sample of included studies to assess clarity, consistency, and relevance. Any modifications made during the extraction process will be documented in the final scoping review.

Discrepancies between reviewers will be resolved through discussion, with a third reviewer adjudicating if needed. Where necessary, study authors will be contacted to obtain missing or additional data. Critical appraisal of individual sources of evidence will not be conducted, consistent with JBI guidance for scoping reviews.

### Data analysis and presentation

**Table 1.** Summary of included studies and change ideas Example extraction table. Example extraction table

| Author | Year | Country | Design | Participants (n) | Facility Type | Barriers and Facilitators | Targeted Domain (SSNC) | Specific Change Idea | Proctors Outcomes Measured (with timeframe and units) | Evidence | Comments |
| --- | --- | --- | --- | --- | --- | --- | --- | --- | --- | --- | --- |
| Article 1 |  |  |  |  |  |  |  |  |  |  |  |
| Article 2 |  |  |  |  |  |  |  |  |  |  |  |

#### Findings

A narrative summary will accompany the tables to describe how strategies, change ideas, and facility-level activities relate to increased adoption of CPAP for small and sick newborns, as well as improvements in CPAP quality. Findings will be organized thematically around key implementation barriers according to the SSNC Core Component Framework.

A comprehensive summary table will report key characteristics for all included studies, including country, facility type, intervention details, and reported outcomes. Where relevant, findings will be mapped to implementation frameworks to identify common contextual factors, adaptations, and strategies. Studies reporting improvements in CPAP coverage, timeliness, or quality will be highlighted.

Consistent with JBI guidance for scoping reviews, no critical appraisal or meta-analysis will be conducted. The synthesis will support evidence-informed decision-making for neonatal CPAP implementation in low-resource settings.

## Data Availability

All data produced in this scoping review comes from pre-published work.

## Acknowledgements

We gratefully acknowledge the contributions of NEST360 for their ongoing efforts to improve the coverage and quality of CPAP for small and sick newborns across sub-Saharan Africa. Their work provided the inspiration and highlighted the need for this literature review and continues to inform global efforts to strengthen newborn care through evidence-based implementation and innovation.

## Declarations

The authors have nothing to declare.

## Author contributions

SM conceptualized the review, served as the primary investigator, and led the manuscript drafting and revisions. KD and CM provided methodological guidance and supported the refinement of the review protocol, data base selection, and search strategy development. SM and NK served as the two reviewers who performed screening and data extraction. KD served as a third reviewer when consensus could not be reached. LH oversaw the review process and provided supervisory input throughout the project. All authors reviewed and approved the manuscript prior to submission for publication.

## Appendices

## Appendix A: Complete inclusion and exclusion criteria OR Inclusion exclusion screening tool

Appendix A : Complete inclusion and exclusion criteria OR Inclusion exclusion screening tool
| Protocol item | A priori criteria |
| --- | --- |
| Review title | Scoping Review CPAP Implementation Strategies for Optimized Neonatal Care |
| Review question | What change ideas and implementation strategies that have been used to promote timely CPAP initiation and use for eligible newborns in clinical settings? |
| Searched | Searches will be completed in three databases: Embase (via Elsevier), PubMed (via NLM), and CINAHL (via EBSCOhost) |
| Types of studies to be included | Inclusion:<br>1) Experimental and quasi-experimental studies (RCTs, non-RCTs, before-and-after, interrupted time series)<br>2) Analytical observational studies (cohort, case-control, analytical cross-sectional)<br>3) Descriptive observational studies (case series, case reports, descriptive cross-sectional)<br>4) Systematic reviews that meet inclusion criteria<br>Exclusion:<br>1) Animal studies<br>2) Text and opinion papers<br>3) Conference abstracts without full text |
| Participant/populations | Inclusion: Small and sick newborns (SSNC), including premature infants, neonates <28 days, low birthweight (<2500g), or those presenting with symptoms of respiratory distress syndrome (RDS).<br>Exclusion: Infants >28 days, non-neonatal populations, neonates without documented RDS risk factors or symptoms |
| Condition or domain being studied | Implementation of CPAP therapy for eligible newborns<br>Inclusion:<br>1) CPAP use for RDS<br>2) CPAP implementation strategies and change ideas<br>Exclusion:<br>1) Studies solely focused on CPAP efficacy<br>2) CPAP use for unrelated conditions (e.g., sleep apnea)<br>3) Studies describing barriers without actionable solutions |
| Context | Clinical settings focused on newborn care<br>Inclusion:<br>1) Neonatal intensive care units<br>2) Standard neonatal units<br>3) Inpatient care environments for at-risk infants<br>4) Delivery rooms and obstetric units if CPAP initiation is emphasized<br>Exclusion:<br>1) Outpatient or community-based settings<br>2) Adult or pediatric ICUs<br>3) Obstetric settings without neonatal CPAP focus |
| Outcomes | Reported implementation strategies, change ideas, quality improvement interventions, barriers and facilitators, and evidence of success or failure in promoting timely CPAP initiation |
| Frameworks used | 1) JBI Scoping Review Methodology<br>2) PRISMA-ScR (Preferred Reporting Items for Systematic Reviews and Meta-Analyses Extension for Scoping Reviews) |

*Note.* Protocol adapted from the PROSPERO systematic review protocol registration form (University of York Centre for Reviews and Dissemination, 2016).

## Appendix B: Search strategy

### 1. Embase (via Elsevier)

**Search conducted on:** August 2025

**Planned limits:** English language only; no date restrictions applied

**Final Search Query:**

(‘newborn’/exp OR ‘full term infant*’:ti,ab OR ‘neonat*’:ti,ab,jt OR ‘newborn*’:ti,ab,jt OR ‘newly born’:ti,ab,jt OR ‘premature infant*’:ti,ab,jt OR ‘preterm*’:ti,ab,jt OR ‘preemie’:ti,ab,jt OR ‘preemies’:ti,ab,jt OR ‘low birth weight’:ti,ab OR ‘low birthweight’:ti,ab OR ‘small for gestational age’:ti,ab OR ‘sga infant*’:ti,ab OR ‘premature birth’:ti,ab OR ‘nicu’:ti,ab OR ‘lbw’:ti,ab OR ‘vlbw’:ti,ab OR ‘neonatologist’/exp OR ‘newborn care’/exp OR ‘newborn mortality’/de OR ‘neonatal intensive care unit’/exp OR ‘low birth weight’/exp OR ‘neonatology’/de OR ‘newborn disease’/exp) AND

(‘continuous positive airway pressure’/exp OR ‘continuous positive airway pressure’:ti OR ‘cpap’:ti OR ‘constant positive airway pressure’:ti,ab OR ‘nasal cpap’:ti,ab OR ‘neonatal cpap’:ti,ab OR ‘ncpap’:ti,ab OR ‘cpap device’/exp) AND

(‘quality improvement’/exp OR ‘guideline adherence’/exp OR ‘training’/exp OR ‘feasibility study’/exp OR ‘implementation science’/exp OR ‘translational research’/exp OR ‘knowledge translation’/exp OR ‘health care planning’/exp OR ‘program evaluation’/exp OR ‘program development’/exp OR ‘quality improvement’:ti,ab,jt OR ‘training’:ti,ab,jt OR ‘implement*’:ti,ab,jt OR ‘feasib*’:ti,ab OR ‘adopt*’:ti,ab OR ‘routine use’:ti,ab OR ‘uptake’:ti,ab OR ‘sustainab*’:ti,ab OR ‘sustained use’:ti,ab OR ‘optimiz*’:ti,ab OR facilitat*:ti,ab OR barrier*:ti,ab)

**Records retrieved:** 1,249 (as of August 18, 2025)

### 2. PubMED (via NLM)

**Search conducted on:** August 2025

**Planned limits:** English language only; no date restrictions applied

**Final Search Query:**

(“Infant, Newborn”[MeSH] OR “Infant, Premature”[MeSH] OR “Infant, Low Birth Weight”[MeSH] OR “Intensive Care Units, Neonatal”[MeSH] OR “Neonatology”[MeSH] OR “Infant Mortality”[MeSH] OR “Infant, Small for Gestational Age”[MeSH] OR “Neonatal Nursing”[MeSH] OR “Infant, Newborn, Diseases”[MeSH] OR “Intensive Care, Neonatal”[MeSH] OR “Neonatologists”[MeSH] OR “Premature Birth”[MeSH] OR “Neonatal Screening”[MeSH] OR “full term infant*”[tiab] OR neonat*[tiab] OR newborn*[tiab] OR “newly born”[tiab] OR “preterm infant*”[tiab] OR “premature infant*”[tiab] OR preterm*[tiab] OR preemie[tiab] OR preemies[tiab] OR “low birth weight”[tiab] OR low birthweight[tiab] OR “small for gestational age”[tiab] OR “sga infant*”[tiab] OR “premature birth”[tiab] OR nicu[tiab] OR lbw[tiab] OR vlbw[tiab])

AND

(“Continuous Positive Airway Pressure”[MeSH] OR “cpap”[tiab] OR “constant positive airway pressure”[tiab] OR “nasal cpap”[tiab] OR “neonatal cpap”[tiab] OR ncpap[tiab] OR “cpap device”[tiab] OR “continuous positive airway pressure”[tiab])

AND

(“Quality Improvement”[MeSH] OR “Guideline Adherence”[MeSH] OR “Feasibility Studies”[MeSH] OR “Implementation Science”[MeSH] OR “Translational Research, Biomedical”[MeSH] OR “Health Planning”[MeSH] OR “Program Evaluation”[MeSH] OR “Program Development”[MeSH] OR “quality improvement”[tiab] OR training[tiab] OR implement*[tiab] OR feasib*[tiab] OR adopt*[tiab] OR “routine use”[tiab] OR uptake[tiab] OR sustainab*[tiab] OR “sustained use”[tiab] OR optimiz*[tiab] OR facilitat*[tiab] OR barrier*[tiab])

**Records retrieved:** 696 (as of August 18, 2025)

### 2. CINAHL (via EBSCOhost)

**Search conducted on:** August 2025

**Planned limits:** English language only; no date restrictions applied

**Final Search Query**

(XB (“full term infant*” OR neonat* OR newborn* OR “newly born” OR “preterm infant*” OR “premature infant*” OR preterm* OR preemie OR preemies OR “low birth weight” OR low birthweight OR “small for gestational age” OR “sga infant*” OR “premature birth” OR nicu OR lbw OR vlbw) OR MH “Infant, Premature”) AND (XB (cpap OR “constant positive airway pressure” OR “nasal cpap” OR “neonatal cpap” OR ncpap) OR MH “Continuous Positive Airway Pressure”) AND (XB (“quality improvement” OR training OR implement* OR feasib* OR adopt* OR “routine use” OR uptake OR sustainab* OR “sustained use” OR optimiz* OR facilitat* OR barrier*) OR MH “Program Implementation”)

**Records retrieved:** 231 (as of August 18, 2025)

**Summary**

|  |  |
| --- | --- |
| Embase | 1249 |
| PubMed | 696 |
| CINAHL | 231 |
| **Total** | **2176** |
| **After removing duplicate records** | **1537** |

## Appendix C: Data extraction instrument

| Domain | Data Field |
| --- | --- |
| Citation Details | Author(s), Year, Title, Journal, Country |
| Study Characteristics | Study design, Setting (Facility/Unit), Sample size, Population described |
| Concept | Intervention type, Implementation strategy, Change idea |
| Context | Country income level, Health system level, Facility type, Neonatal service tier |
| SSNC Component(s) | Select all that apply:<br>• Leadership & Governance<br>• Financing<br>• Infrastructure<br>• Equipment & Commodities<br>• Data Systems & QI<br>• Human Resources<br>• Referral System<br>• Linkage to Maternal Care<br>• Post-Discharge Follow-Up |
| Proctors Implementation Outcomes<br>(including timeframe and units) | Acceptability, Adoption, Appropriateness, Feasibility, Fidelity, Implementation Cost, Penetration, Sustainability. |
| Barriers & Facilitators | Reported challenges and enabling factors |
| Evidence/Key Findings | Summary of results relevant to CPAP implementation or neonatal care |
| Notes | Reviewer comments, uncertainties, follow-up needed |

